# The heterogeneity of youth at risk of diabetes and prediabetes: a latent class analysis of a national sample

**DOI:** 10.1101/2024.11.20.24317640

**Authors:** Catherine McDonough, Yan Chak Li, Gaurav Pandey, Nita Vangeepuram, Bian Liu

**Affiliations:** Department of Population Health Science and Policy, Icahn School of Medicine at Mount Sinai, New York, NY, USA; Department of Genetics and Genomic Sciences, Icahn School of Medicine at Mount Sinai, New York, NY, USA; Department of Pediatrics, Icahn School of Medicine at Mount Sinai, New York, NY, USA; Institute for Health Equity Research, Icahn School of Medicine at Mount Sinai, New York, NY, USA

**Author notes:** Corresponding author Bian Liu, Department of Population Health Science and Policy, Icahn School of Medicine at Mount Sinai, One Gustave L. Levy Place, Box 1077, New York, NY, USA 10029.

## Abstract

**Objectives:** To identify youth subgroups based on lifestyle, BMI, and sociodemographic characteristics and examine the association between group membership and prediabetes/diabetes (preDM/DM) status.

**Methods:** We analyzed data from 1,278 adolescents (ages 12-17) from the 2011-2018 National Health and Nutrition Examination Surveys. PreDM/DM was defined using hemoglobin A1c (≥5.7 mg/dL) and/or fasting plasma glucose (≥100 mg/dL). Latent class analysis of physical activity, diet quality, screen time, and BMI identified subgroups, adjusted for sociodemographic factors. Associations between class membership and preDM/DM were assessed using survey-weighted logistic regression.

**Results:** Four classes emerged: High BMI and unhealthy lifestyle (37.5%), Healthy BMI and physically active (25.3%), Healthy BMI and lifestyle (16.0%), and Average BMI and lifestyle (21.8%). Youth in other classes had lower odds of preDM/DM compared to the High BMI and unhealthy lifestyle class, especially the Healthy BMI and active class (aOR=0.556, 95% CI=0.327-0.946).

**Conclusions:** Youth at risk of preDM/DM were from heterogeneous groups with varied lifestyle, health, and socioeconomic characteristics.

## Introduction

Type 2 diabetes mellitus (DM) and prediabetes (preDM) are multifaceted conditions influenced by various biological^1,2^ and epidemiological factors^1,3,4^, dietary patterns^2,5^, physical activity^2,6–8^, and socioeconomic status^9,10^. Recent data reveal a surge in preDM/DM prevalence among youth in the United States from 4.1% in 1999 to 22% in 2018^11^, and the trend is only projected to continue^12,13^. This is especially troubling due to its disproportionate impact on racial and ethnic minority groups and those with limited socioeconomic resources, exacerbating existing health disparities^9,10^. Early onset of preDM/DM poses heightened health and economic burdens due to prolonged disease duration and increased susceptibility to other cardiometabolic conditions^14–16^. Intervening in this pressing issue requires intensified research into the interplay of multiple preDM/DM risk factors to inform better prevention strategies.

There is a large body of research addressing individual preDM/DM risk factors, such as physical activity, body mass index (BMI), diet, and screen time among youth. Notably, studies have shown strong associations between increased physical activity and reduced risk of preDM/DM as well as benefits in managing these conditions once developed^6,7^. Higher BMI and unhealthy dietary patterns have emerged as significant indicators of preDM/DM susceptibility^1,4,5^. The impact of screen time on preDM/DM risk remains unclear, with mixed findings in the literature. While most studies indicate a potential correlation between excessive screen time and preDM/DM risk^17–20^, others have found inconclusive or contradictory evidence^21^. These studies, while useful, also have limitations by examining risk factors individually, despite recognition that multiple factors likely work together to affect the risk of youth preDM/DM.

To address this gap, we applied latent class analysis (LCA) to a nationally representative sample of youth with data on physical activity, diet, screen time, BMI, and sociodemographic factors. LCA is a statistical method that categorizes individuals into latent groups with similar observable characteristics^22^. LCA is, thus, advantageous to identify underlying subpopulations based on preDM/DM risk factors.

The objectives of the study were to identify distinct subgroups characterized by unique combinations of lifestyle factors and BMI using LCA and to examine the association between the latent class membership and preDM/DM status. Discerning unique combinations of potential preDM/DM risk factors may enhance our understanding of preDM/DM etiology and facilitate targeted approaches for high-risk subgroups.

## Methods

### Study Population

This study utilized publicly available data from the National Health and Nutrition Examination Survey (NHANES), a nationally representative survey conducted by the Center for Disease Control (CDC)^23^. We leveraged our pre-processed NHANES data from the Prediabetes/diabetes in youth ONline Dashboard (POND)^11^ and restricted the sample with additional requirements: (i) interviewed during cycles from 2011 to 2018, (ii) aged between 12 and 17 years, (iii) had non-zero and non-missing fasting plasma glucose survey weight, and (iv) had complete data for variables of interest, for a final sample of 1,278 youth (Figure S1).

### PreDM/DM outcome

We determined preDM/DM status based on criteria outlined by the American Diabetes Association, where individuals with hemoglobin A1c of 5.7 mg/dL or greater, and/or fasting plasma glucose of 100 mg/dL or greater were considered to be at risk of preDM/DM^1^.

#### Body Mass Index (BMI)

BMI percentiles were calculated from weight and height utilizing the SAS Program from the CDC^24^, using the 2000 CDC BMI-for-Age Growth Charts. The percentiles were categorized in accordance with established pediatric percentile cut-offs for the child’s specific age and sex. We grouped BMI percentiles into three categories: underweight or normal (<85th%ile), overweight (85th to 95th%ile), and obese (>95th%ile). Due to the small sample size (n=25, 2.0% of the analytical sample) for the underweight group (BMI percentile<5th%ile), the underweight and normal weight youth were collapsed into one group.

#### Physical activity

We assessed physical activity as a composite variable based on data availability through the NHANES cycles^23^. From 2011-2016, physical activity was defined as self-reported moderate or vigorous activity in hours per week. Moderate activities, such as walking, result in slight increases in heart rate, while vigorous ones, such as running, lead to more significant increases^25^. Participants reported the number of days and average minutes per day of each activity. We then calculated average daily hours. For 2017-2018, an alternative question asked the number of days per week participants were physically active for at least 60 minutes. We estimated that each day corresponded to 60 minutes of physical activity, recognizing that this approximation may underestimate the time for some (Table S1). The final physical activity variable was categorized into survey-weighted quartiles with cut-offs at 0.18, 0.56, and 0.99 hours per day.

#### Diet quality

NHANES collects data on food eaten in the last 24 hours from two assessments^26–28^. We utilized data from the first assessment to calculate the Health Eating Index (HEI) score, which measures how well participants’ diets align with the United States Department of Agriculture (USDA) dietary guidelines^29^. HEI scores range from 0 to 100. We calculated the overall HEI score (HEI-2015) using SAS code provided by the USDA compatible with the NHANES Total Nutrient (First Day) variables^30^. HEI scores were categorized into survey-weighted quartiles with cut-offs at 35.70, 44.77, and 53.51.

#### Screen time

Screen time was estimated by self-reported average hours per day watching television or using computers outside of school based on two questions. The first question asked average daily hours spent watching television or videos over the preceding 30 days, and the second question asked the same but of computer usage. The summation of these two components yielded the total screen time hours per day. Screen time was categorized into survey-weighted quartiles with cut-offs at 2.01, 4.17, and 7.20 hours per day.

### Sociodemographic covariates

Given the importance of sociodemographic factors on preDM/DM risk^9^, we included the following covariates: sex, race/ethnicity, family poverty income ratio (PIR), and insurance status. Family PIR is defined as the ratio of family income to the federal poverty level based on family size and state of residence^31^, with value below 1 indicating the family income is below the poverty line. PIR was categorized into four categories. The first category comprises any PIR value below 1. The three subsequent categories pertain to tertiles for the remaining sample. We categorized insurance status into four groups: private, government (excluding Medicaid and Children’s Health Insurance Program (CHIP)), Medicaid/CHIP, and no insurance.

### Statistical Analysis

Unless otherwise specified, all analyses accounted for NHANES complex survey design, using fasting glucose subsample weight^32^.

We examined descriptive characteristics of the overall sample and by preDM/DM status and latent class membership using median and interquartile range for continuous variables and frequencies and percentiles for categorical variables. We compared characteristics across subgroups using survey-weighted linear regression for continuous variables and Rao-Scott Chi Square tests for categorical variables.

We conducted LCA using the poLCA package in R (version 4.2.2; R Core Team, 2022) to identify distinct lifestyle and BMI subgroups. We tested unweighted LCA models including 2 to 6 classes, while adjusting for sex, race/ethnicity, family PIR, and insurance status. We determined the optimal number of latent classes based on the lowest median Akaike information criterion (AIC) determined from a bootstrap analysis of 100 runs, sampling with replacement, as well as reasonable class sizes and interpretability. Once the optimal number of classes was determined, we assessed the association between latent class membership (the exposure variable) and the risk of preDM/DM (the outcome variable) using a survey-weighted logistic regression model, while adjusting for the aforementioned covariates. To account for the uncertainty of the class membership assignment, we also conducted a sensitivity analysis using the Bolck, Croon, and Hagenaars (2004) (BCH) three-step approach^33^.

## Results

### Study population characteristics

Table 1 displays the survey-weighted characteristics overall and by preDM/DM status. Both groups showed similar distribution in terms of median age in years (14.2 and 14 for youth with and without preDM/DM). The racial-ethnic distribution was also comparable between the groups, with the majority being Non-Hispanic White (55.1% for preDM/DM and 53.2% for non-preDM/DM). Economic characteristics, as indicated by family PIR and health insurance status, were similar between the groups. Youth with preDM/DM had a higher proportion of males (66.2% vs. 41.8%, p-value<0.0001) and a higher prevalence of obesity (34.2% vs. 19.0% in the non-preDM/DM group). Additionally, we observed a notable increase in prevalence across survey cycles, from 23.3% to 41.6%.

**Table 1.** Survey weighted study population characteristics by preDM/DM status, NHANES 2011-2018.

|  | Overall<br>(n=1,278) | Without preDM/DM<br>(n=878,<br>wgt % = 68.7) | With preDM/DM<br>(n=400,<br>wgt % = 31.3) | P-Value |
| --- | --- | --- | --- | --- |
| **Age (years)** | 14.2 (12.6, 15.5) | 14.2 (12.6, 15.6) | 14 (12.7, 15.4) | 0.397 |
| **Sex** |  |  |  | <0.0001 |
| Male | 619 (49.4) | 365 (41.8) | 254 (66.2) |  |
| Female | 659 (51.6) | 513 (58.2) | 146 (33.8) |  |
| **Race/ethnicity** |  |  |  | 0.711 |
| Hispanic | 392 (22.1) | 276 (22.4) | 116 (21.5) |  |
| Non-Hispanic White | 367 (54.5) | 250 (55.1) | 117 (53.2) |  |
| Non-Hispanic Black | 312 (13.9) | 209 (13) | 103 (15.8) |  |
| Other | 207 (9.6) | 143 (9.6) | 64 (9.6) |  |
| **Family Poverty Income Ratio (PIR)** |  |  |  | 0.514 |
| Below poverty (PIR <= 1) | 393 (21.6) | 270 (21.1) | 123 (22.6) |  |
| 1st tertile (1 < PIR < 1.85) | 337 (22.5) | 228 (22.6) | 109 (22.1) |  |
| 2nd tertile (1.85 <= PIR < 3.60) | 309 (30.3) | 207 (29) | 102 (33.3) |  |
| 3rd tertile (3.60 <= PIR) | 239 (25.6) | 173 (27.3) | 66 (22) |  |
| **Insurance status** |  |  |  | 0.537 |
| Private | 473 (44.9) | 329 (44.8) | 144 (45.1) |  |
| Government | 215 (19) | 146 (19.9) | 69 (17) |  |
| Medicaid/CHIP | 469 (28.1) | 317 (26.8) | 152 (30.9) |  |
| No insurance | 121 (8) | 86 (8.5) | 35 (6.9) |  |
| **Body Mass Index (BMI)** |  |  |  | 0.0001 |
| Underweight or normal weight (BMI %ile < 85th) | 736 (58.4) | 540 (62.4) | 196 (49.6) |  |
| Overweight (85th BMI %ile < 95th) | 217 (17.8) | 160 (18.6) | 57 (16.2) |  |
| Obese (95th ≤ BMI %ile) | 325 (23.8) | 178 (19) | 147 (34.2) |  |
| **Healthy Eating Index (HEI)** |  |  |  | 0.082 |
| 1st quartile (HEI < 35.70) | 309 (25.4) | 202 (23.8) | 107 (28.8) |  |
| 2nd quartile (35.70 ≤ HEI < 44.77) | 326 (24.5) | 226 (25) | 100 (23.4) |  |
| 3rd quartile (44.77 ≤ HEI < 53.51) | 319 (24.9) | 213 (23.6) | 106 (28) |  |
| 4th quartile (53.51 ≤ HEI) | 324 (25.2) | 237 (27.6) | 87 (19.8) |  |
| **Physical activity hours per day** |  |  |  | 0.395 |
| 1st quartile (hours < 0.18) | 350 (24.3) | 249 (25.8) | 101 (21) |  |
| 2nd quartile (0.18 ≤ hours < 0.56) | 315 (24) | 211 (23) | 104 (26.4) |  |
| 3rd quartile (0.56 ≤ hours < 0.99) | 286 (23.9) | 187 (22.9) | 99 (26) |  |
| 4th quartile (0.99 ≤ hours) | 327 (27.8) | 231 (28.3) | 96 (26.6) |  |
| **Screen time hours per day** |  |  |  | 0.222 |
| 1st quartile (hours < 2.01) | 302 (25.1) | 216 (25.9) | 86 (23.3) |  |
| 2nd quartile (2.01 ≤ hours < 4.17) | 310 (23.5) | 224 (25.1) | 86 (20) |  |
| 3rd quartile (4.17 ≤ hours < 7.20) | 325 (25.4) | 214 (23.7) | 111 (29) |  |
| 4th quartile (7.20 ≤ hours) | 341 (26) | 224 (25.2) | 117 (27.6) |  |
| **Survey Cycle** |  |  |  | 0.0007 |
| 2011-2012 | 353 (26.6)<sup>a</sup> | 264 (76.8) | 89 (23.2) |  |
| 2013-2014 | 351 (23.9) | 265 (75.2) | 86 (24.8) |  |
| 2015-2016 | 307 (25.9) | 196 (63.9) | 111 (36.1) |  |
| 2017-2018 | 267 (23.5) | 153 (58.4) | 114 (41.6) |  |

<sup>a</sup>Percentages for survey cycle are presented as column percents rather than row percents for more meaningful interpretation purposes.

### Latent classes by BMI and lifestyle variables

We identified that four latent classes best delineated distinct health and activity profiles among our sample: High BMI and unhealthy lifestyle (n=479 (37.5%)), Healthy BMI and physically active (n=316 (25.3%)), Healthy BMI and lifestyle (n=205 (16.0%)), and High BMI and average lifestyle (n=278 (21.8%)) (Figure S2). The class names were based on estimated class-conditional response probabilities (Fig. 1) and were corroborated by the distributions in the sample (Table 2). For instance, youth in the two “high BMI” classes had 0.34 and 0.31 probabilities of obesity and 0.47 and 0.5 probabilities of normal weight. In contrast, youth in the two “healthy BMI” classes had 0.78 and 0.67 probabilities of being normal weight and only 0.11 and 0.16 probabilities of obesity.

**Table 2.**
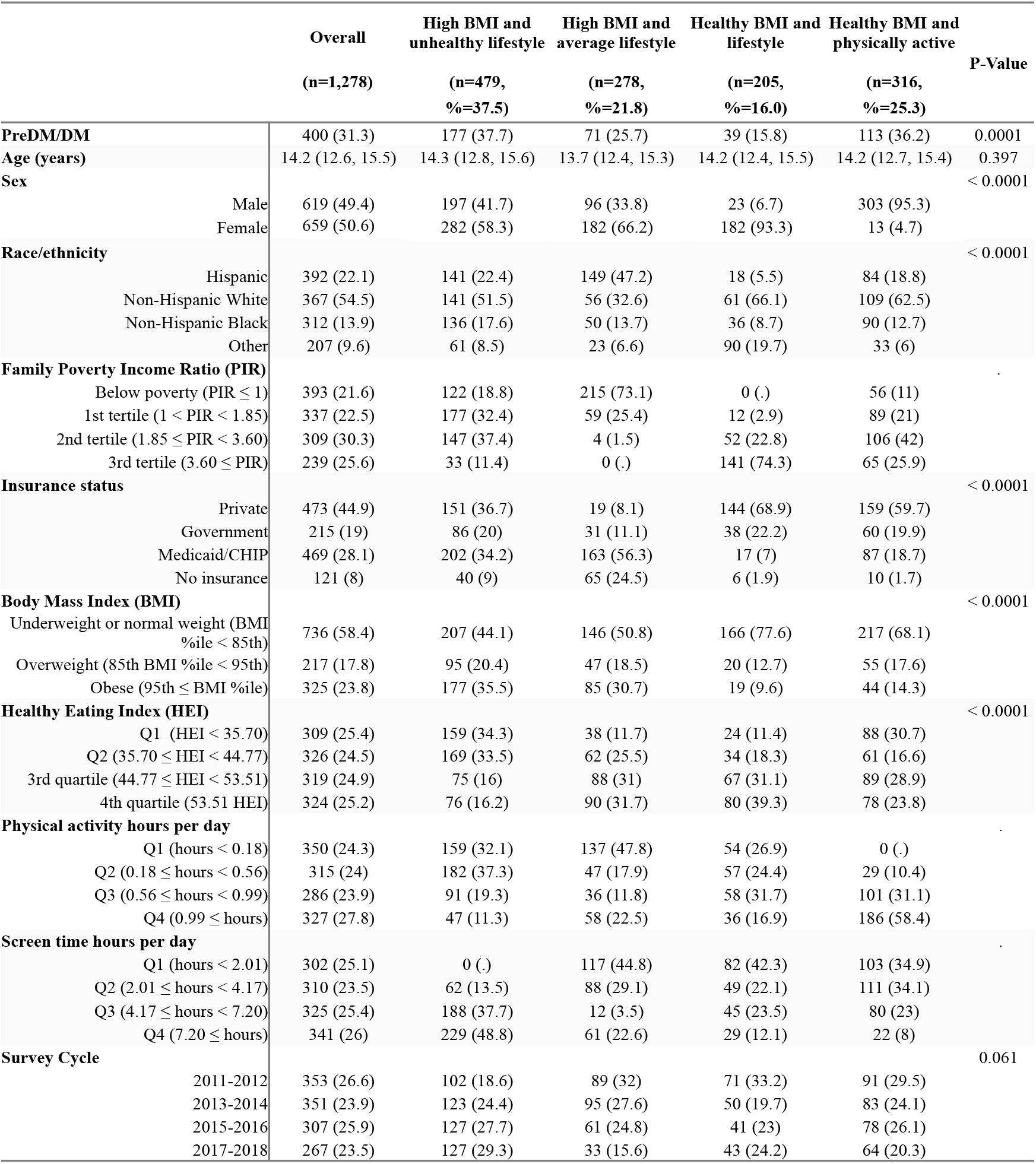
Survey weighted study population characteristics by the identified four latent classes, NHANES 2011-2018.

**Figure 1.**
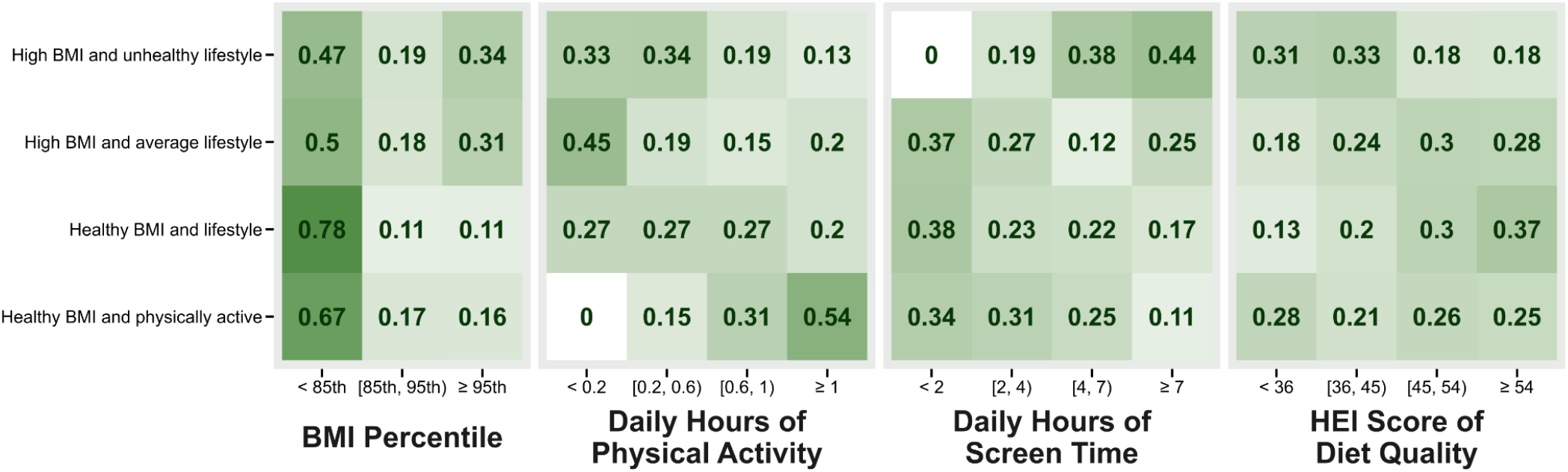
Class-specific probabilities of BMI and lifestyle behavior categories in the four identified latent classes.

Accordingly, the obesity prevalence shown in Table 2 was above 30% for the two “high BMI” classes while below 15% for the two “healthy BMI” classes.

Youth in the “unhealthy lifestyle” class had 0.64 and 0.67 probabilities of below median diet quality and physical activity, respectively. They also had a 0.44 probability of having the highest screen time and zero probability of the lowest screen time (Fig. 1). Descriptively in Table 2, 48.8% of youth in this group had the highest quartile of screen time, compared to less than 23% in the other three latent classes.

Youth in the “Healthy BMI and physically active” class distinguished themselves as the most active class with a 0.54 probability of being in the highest physical activity quartile. In Table 2, 58.4% of youth in the “physically active” class fell into the highest quartile of activity, while the proportion ranged from 11.3% to 27.8% in the other three classes.

### Latent classes by sociodemographic variables

The distribution of covariates also varied based on class membership assignment (Table 2). “Physically active” youth predominantly had private insurance (59.7%) and fell in the 2nd tertile of family PIR (42%). Amongst those with a “Healthy BMI and lifestyle”, 74.3% of the youth fell in the highest tertile of the poverty income ratio (PIR), and 68.9% were covered by private insurance. In terms of racial and ethnic composition, those in the two “Healthy BMI” classes were more likely to identify as Non-Hispanic White (62.5% and 66.1%, respectively). Those in the “Healthy BMI and lifestyle” class were least likely to identify as being from a racially/ethnically minoritized background.

In contrast, the “High BMI and unhealthy lifestyle” class contained the highest proportion of Non-Hispanic Black youth (17.6%) and moderate socioeconomic characteristics with 34.2% covered by Medicaid or CHIP and 69.8% falling in the first and second PIR tertiles. The “High BMI and average lifestyle” class was composed primarily of racial and ethnic minorities with 47.2 % Hispanic and 13.7% non-Hispanic Black youth. This class also had the lowest socioeconomic status of the four classes; 73.1% of youth fell below the poverty line, and 56.3% were covered by Medicaid or CHIP.

It is important to note the enormous sex differences among classes. Those in the “Healthy BMI and physically active” class contained predominantly male youth (95.3%, Table 2). This differs greatly from the class with a “Healthy BMI and lifestyle” which included only 6.7% male youth.

### Association between class membership and youth preDM/DM status

Using youth in the “High BMI and unhealthy lifestyle” class as the reference, we found reduced odds of preDM/DM risk in the other three classes, both before and after adjusting for sociodemographics and survey cycles (Table 3). In the unadjusted model, significant associations were observed for those with a “Healthy BMI and lifestyle” (OR = 0.309, 95% CI [0.167-0.572]) and for those with a “High BMI and average lifestyle” (OR = 0.570, 95% CI [0.372-0.872]), but not for those in the “Healthy BMI and physically active” class (OR = 0.936, 95% CI [0.590-1.486]). In the adjusted model, a significant association was only found for youth in the “Healthy BMI and physically active” class (aOR = 0.556, 95% CI [0.327-0.946]). The associations were not significant in the “Healthy BMI and lifestyle” (aOR = 0.459, 95% CI [0.209-1.008]) and the “High BMI and average lifestyle” classes (aOR = 0.676, 95% CI [0.417-1.097]).

**Table 3.**
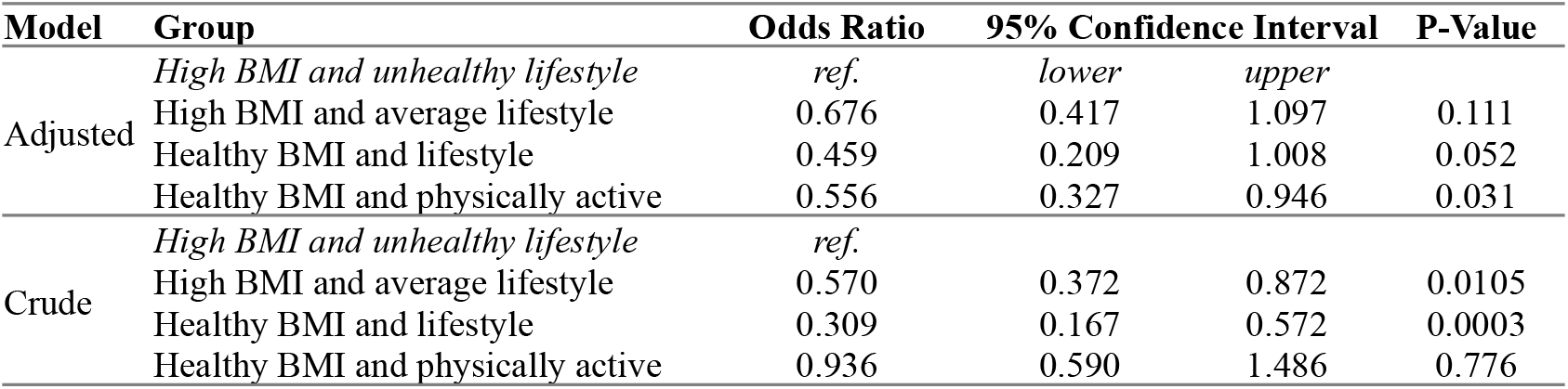
Association between preDM/DM and identified four latent classes.

| **Model** | **Group** | **Odds Ratio** | **95% Confidence Interval** |  | **P-Value** |
| --- | --- | --- | --- | --- | --- |
| Adjusted | *High BMI and unhealthy lifestyle* | *ref.* | *lower* | *upper* |  |
|  | High BMI and average lifestyle | 0.676 | 0.417 | 1.097 | 0.111 |
|  | Healthy BMI and lifestyle | 0.459 | 0.209 | 1.008 | 0.052 |
|  | Healthy BMI and physically active | 0.556 | 0.327 | 0.946 | 0.031 |
| Crude | *High BMI and unhealthy lifestyle* | *ref.* |  |  |  |
|  | High BMI and average lifestyle | 0.570 | 0.372 | 0.872 | 0.0105 |
|  | Healthy BMI and lifestyle | 0.309 | 0.167 | 0.572 | 0.0003 |
|  | Healthy BMI and physically active | 0.936 | 0.590 | 1.486 | 0.776 |

Similar results were found using the 3-step BCH-approach (Figure S3), where we found four comparable classes. Youth with preDM/DM risk had significantly lower odds of belonging to the “Healthy BMI and lifestyle” (aOR=0.38, 95% CI [0.18-0.83]), “Healthy BMI and physically active” (aOR=0.56, 95% CI [0.27-1.18]), and “High BMI and average lifestyle” (aOR=0.46, 95% CI [0.22-0.97]) classes.

## Discussion

Applying latent class analysis to a nationally representative sample, we found youth aged 12-17 years can be grouped into four distinct classes based on their BMI, physical activity, diet, screen time, and sociodemographic factors. More importantly, youth in these four groups also differed in their odds of having preDM/DM. Youth in the “High BMI and unhealthy lifestyle habits” class were most likely to have preDM/DM, indicating that co-occurrence of high BMI and poor lifestyle habits may carry the highest risk of preDM/DM, while low risk of preDM/DM may present in heterogeneous combinations of BMI and lifestyle characteristics. The interplay between lifestyle behaviors, BMI, sociodemographic factors and preDM/DM risk underscores the complex nature of the disease and the need for comprehensive and tailored preventive strategies.

Classes were categorized based on their distribution within this population, but when compared with national recommendations, even our “healthiest” categories fell short. Youth aged 6-17 should engage in at least 1 hour per day of moderate to vigorous exercise^18^, but only 27.8% of youth in this study met this recommendation. Diet quality was poor in this study with only our highest quartile having an HEI score of 54 out of 100, with 100 representing meeting all existing recommendations. The amounts of screen time seen in this study were extremely high, with the lowest quartile at 2 hours per day and the highest over 7 hours, without considering additional time spent using the computer or electronics for educational purposes.

Considering that youth generally did not meet standard health recommendations, it is unsurprising that regardless of class, the prevalence of preDM/DM remained strikingly high, ranging from 15.8% to 37.7%. The potential physical, economic, and psychological burden of DM can be substantial, and having preDM at such an early age increases health and economic burden long-term^16^.

It is well-documented that racial and ethnic minorities and low-income families suffer poorer health outcomes and face higher risks of preDM/DM^9,10^. Two thirds of the youth in the “healthy BMI” categories identified as Non-Hispanic White. Meanwhile those in the “high BMI” categories had higher proportions of Hispanic and Non-Hispanic Black identifying youth, and nearly three quarters of the youth in the “High BMI and average lifestyle” class were from households that fell below the poverty line, and nearly a quarter were not covered by insurance at all. Potential explanations for these between class differences are likely multifactorial and may include limited access to healthier lifestyle choices and healthcare and increased disease burden stemming from perpetuating cycles of disadvantage and limited opportunities for socioeconomic advancement^9,10^.

Consistent with the high risk of preDM/DM among male youth reported in the literature^34^, we found that the “Healthy BMI and physically active” class was almost entirely male (95.3%) but had the second-highest prevalence of preDM/DM. This contrasted sharply with the closely related “Healthy BMI and lifestyle” class, which had the lowest preDM/DM prevalence among the four classes and only 7% males. Despite being more active, the “physically active” class had a substantially greater risk of preDM/DM, likely driven by the predominance of male sex. This suggests that to reduce preDM/DM among boys, efforts focusing on factors other than physical activity might be more effective.

Our study has a few limitations. The cross-sectional nature of NHANES data limited the ability to establish causality. Many of the variables included were based on self-reported data, which are subject to recall and response bias. The questionnaire wording changes across survey cycles introduced some limitations in the estimate of physical activity. Screen time only captured the use of TV and computers but not smartphones or other electronic screens, which have increasingly become the dominant mode of screen time among youth in particular^35^. Future research should utilize longitudinal designs and objective measures of lifestyle behaviors to validate our findings.

## Conclusion

Our study revealed subgroups of youth with different lifestyle, health, and socioeconomic characteristics and their associated preDM/DM risk. The identification of these heterogeneous subgroups may reflect the complexity of the multifactorial nature of preDM/DM. Our study also reinforces the importance of adopting healthy lifestyle behaviors, including regular physical activity, limited screen time, and healthy dietary practices, in preventing preDM/DM, but suggests that strategies may need to be tailored for different groups. Findings highlight the potential of targeted interventions to reduce preDM/DM risk, with consideration of which combinations of lifestyle, BMI, and socioeconomic factors may be most important for specific groups.

## Data Availability

The preprocessed data used in this study are available here: https://rstudio-connect.hpc.mssm.edu/POND/#section-download. The original NHANES dataset is publicly available.

## References

1. ElSayed NA, Aleppo G, Aroda VR, et al. 2. Classification and Diagnosis of Diabetes: Standards of Care in Diabetes—2023. Diabetes Care. 2023;46(Suppl 1):S19–S40.doi:10.2337/dc23-S002

2. Temneanu OR, Trandafir LM, Purcarea MR. Type 2 diabetes mellitus in children and adolescents: a relatively new clinical problem within pediatric practice. J Med Life. 2016;9(3):235–239.

3. Chu P, Patel A, Helgeson V, Goldschmidt AB, Ray MK, Vajravelu ME. Perception and Awareness of Diabetes Risk and Reported Risk-Reducing Behaviors in Adolescents. JAMA Netw Open. 2023;6(5):e2311466.doi:10.1001/jamanetworkopen.2023.11466

4. Cioana M, Deng J, Nadarajah A, et al. The Prevalence of Obesity Among Children With Type 2 Diabetes: A Systematic Review and Meta-analysis. JAMA Netw Open. 2022;5(12):e2247186.doi:10.1001/jamanetworkopen.2022.47186

5. Zhuang P, Liu X, Li Y, et al. Effect of Diet Quality and Genetic Predisposition on Hemoglobin A1c and Type 2 Diabetes Risk: Gene-Diet Interaction Analysis of 357,419 Individuals. Diabetes Care. 2021;44(11):2470–2479.doi:10.2337/dc21-1051

6. Rietz M, Lehr A, Mino E, et al. Physical Activity and Risk of Major Diabetes-Related Complications in Individuals With Diabetes: A Systematic Review and Meta-Analysis of Observational Studies. Diabetes Care. 2022;45(12):3101–3111.doi:10.2337/dc22-0886

7. Colberg SR, Sigal RJ, Yardley JE, et al. Physical Activity/Exercise and Diabetes: A Position Statement of the American Diabetes Association. Diabetes Care. 2016;39(11):2065–2079.doi:10.2337/dc16-1728

8. Pivovarov JA, Taplin CE, Riddell MC. Current perspectives on physical activity and exercise for youth with diabetes: Perspectives on exercise. Pediatr Diabetes. 2015;16(4):242–255.doi:10.1111/pedi.12272

9. Hill-Briggs F, Adler NE, Berkowitz SA, et al. Social Determinants of Health and Diabetes: A Scientific Review. Diabetes Care. 2021;44(1):258–279.doi:10.2337/dci20-0053

10. Butler AM. Social Determinants of Health and Racial/Ethnic Disparities in Type 2 Diabetes in Youth. Curr Diab Rep. 2017;17(8):60.doi:10.1007/s11892-017-0885-0

11. McDonough C, Li YC, Vangeepuram N, Liu B, Pandey G. A Comprehensive Youth Diabetes Epidemiological Data Set and Web Portal: Resource Development and Case Studies. JMIR Public Health Surveill. 2024;10(1):e53330.doi:10.2196/53330

12. Tönnies T, Brinks R, Isom S, et al. Projections of Type 1 and Type 2 Diabetes Burden in the US Population Aged <20 Years through 2060: The SEARCH for Diabetes in Youth Study.; 2022.doi:10.2337/figshare.21514014

13. Imperatore G, Boyle JP, Thompson TJ, et al. Projections of type 1 and type 2 diabetes burden in the U.S. population aged <20 years through 2050: dynamic modeling of incidence, mortality, and population growth. Diabetes Care. 2012;35(12):2515–2520.doi:10.2337/dc12-0669

14. O’Connell JM, Manson SM. Understanding the Economic Costs of Diabetes and Prediabetes and What We May Learn About Reducing the Health and Economic Burden of These Conditions. Diabetes Care. 2019;42(9):1609–1611.doi:10.2337/dci19-0017

15. Stedman M, Lunt M, Davies M, et al. Cost of hospital treatment of type 1 diabetes (T1DM) and type 2 diabetes (T2DM) compared to the non-diabetes population: a detailed economic evaluation. BMJ Open. 2020;10(5):e033231.doi:10.1136/bmjopen-2019-033231

16. American Diabetes Association. Economic Costs of Diabetes in the U.S. in 2017. Diabetes Care. 2018;41(5):917–928.doi:10.2337/dci18-0007

17. Kanaley JA, Colberg SR, Corcoran MH, et al. Exercise/Physical Activity in Individuals with Type 2 Diabetes: A Consensus Statement from the American College of Sports Medicine. Med Sci Sports Exerc. 2022;54(2):353–368.doi:10.1249/MSS.0000000000002800

18. Katzmarzyk PT, Powell KE, Jakicic JM, Troiano RP, Piercy K, Tennant B. Sedentary Behavior and Health: Update from the 2018 Physical Activity Guidelines Advisory Committee. Med Sci Sports Exerc. 2019;51(6):12271241.doi:10.1249/MSS.0000000000001935

19. Zheng Y, Ley SH, Hu FB. Global aetiology and epidemiology of type 2 diabetes mellitus and its complications. Nat Rev Endocrinol. 2018;14(2):88–98.doi:10.1038/nrendo.2017.151

20. van der Berg JD, Stehouwer CDA, Bosma H, et al. Associations of total amount and patterns of sedentary behaviour with type 2 diabetes and the metabolic syndrome: The Maastricht Study. Diabetologia. 2016;59(4):709–718.doi:10.1007/s00125-015-3861-8

21. Ekelund U, Brage S, Griffin SJ, Wareham NJ. Objectively Measured Moderate- and Vigorous-Intensity Physical Activity but Not Sedentary Time Predicts Insulin Resistance in High-Risk Individuals. Diabetes Care. 2009;32(6):1081–1086.doi:10.2337/dc08-1895

22. Samuelsen KM, Dayton CM. Latent class analysis. In: The Reviewer’s Guide to Quantitative Methods in the Social Sciences, 2nd Ed. Routledge/Taylor & Francis Group; 2019:164–177.doi:10.4324/9781315755649-12

23. Centers for Disease Control and Prevention (CDC). National Health and Nutrition Examination Survey Data.

24. The SAS Program for CDC Growth Charts. https://www.cdc.gov/nccdphp/dnpao/growthcharts/resources/sas.htm

25. PAQ_I. Accessed May 14, 2024. https://wwwn.cdc.gov/Nchs/Nhanes/2015-2016/PAQ_I.htm

26. Zipf G, Chiappa M, Porter KS, Ostchega Y, Lewis BG, Dostal J. National health and nutrition examination survey: plan and operations, 1999-2010. Vital Health Stat Ser 1 Programs Collect Proced. 2013;(56):1–37.

27. Food Surveys Research Group, Health USDO, Control CFD, National Center For Health Statistics. What We Eat In America (WWEIA). Published online 2015.doi:10.15482/USDA.ADC/1178144

28. NHANES - Measuring Guides. May 8, 2019. Accessed February 29, 2024. https://www.cdc.gov/nchs/nhanes/measuring_guides_dri/measuringguides.htm

29. Healthy Eating Index (HEI) | Food and Nutrition Service. Accessed February 29, 2024. https://www.fns.usda.gov/cnpp/healthy-eating-index-hei

30. Healthy Eating Index SAS Code | EGRP/DCCPS/NCI/NIH. Accessed February 29, 2024. https://epi.grants.cancer.gov/hei/sas-code.html Bureau UC. How the Census

31. Bureau Measures Poverty. Census.gov. Accessed February 29, 2024. https://www.census.gov/topics/income-poverty/poverty/guidance/poverty-measures.html

32. Vital and Health Statistics, Series 2, Number 184.

33. The Methodology Center, Penn State. Accessed Latent Class Analysis (LCA) Covariates 3-Step SAS Macro.doi:10.26207/XP5X-5F64

34. Liu J, Li Y, Zhang D, Yi SS, Liu J. Trends in Prediabetes Among Youths in the US From 1999 Through 2018. JAMA Pediatr. 2022;176(6):608–611.doi:10.1001/jamapediatrics.2022.0077

35. Arundell L, Parker K, Timperio A, Salmon J, Veitch J. Home-based screen time behaviors amongst youth and their parents: familial typologies and their modifiable correlates. BMC Public Health. 2020;20(1):1492.doi:10.1186/s12889-020-09581-w

