## Supplementary Materials for "The heterogeneity of youth at risk of diabetes and prediabetes: a latent class analysis of a national sample"

**The PDF file includes:**
Figure S1: Patient selection flow chart
Figure S2: Bootstrap class selection by AIC
Figure S3: Bolck, Croon, and Hagenaars method latent class probabilities
Table S1: Raw NHANES variables and corresponding cycles used to build physical activity composite variable

**Figure S1. Patient selection criteria**

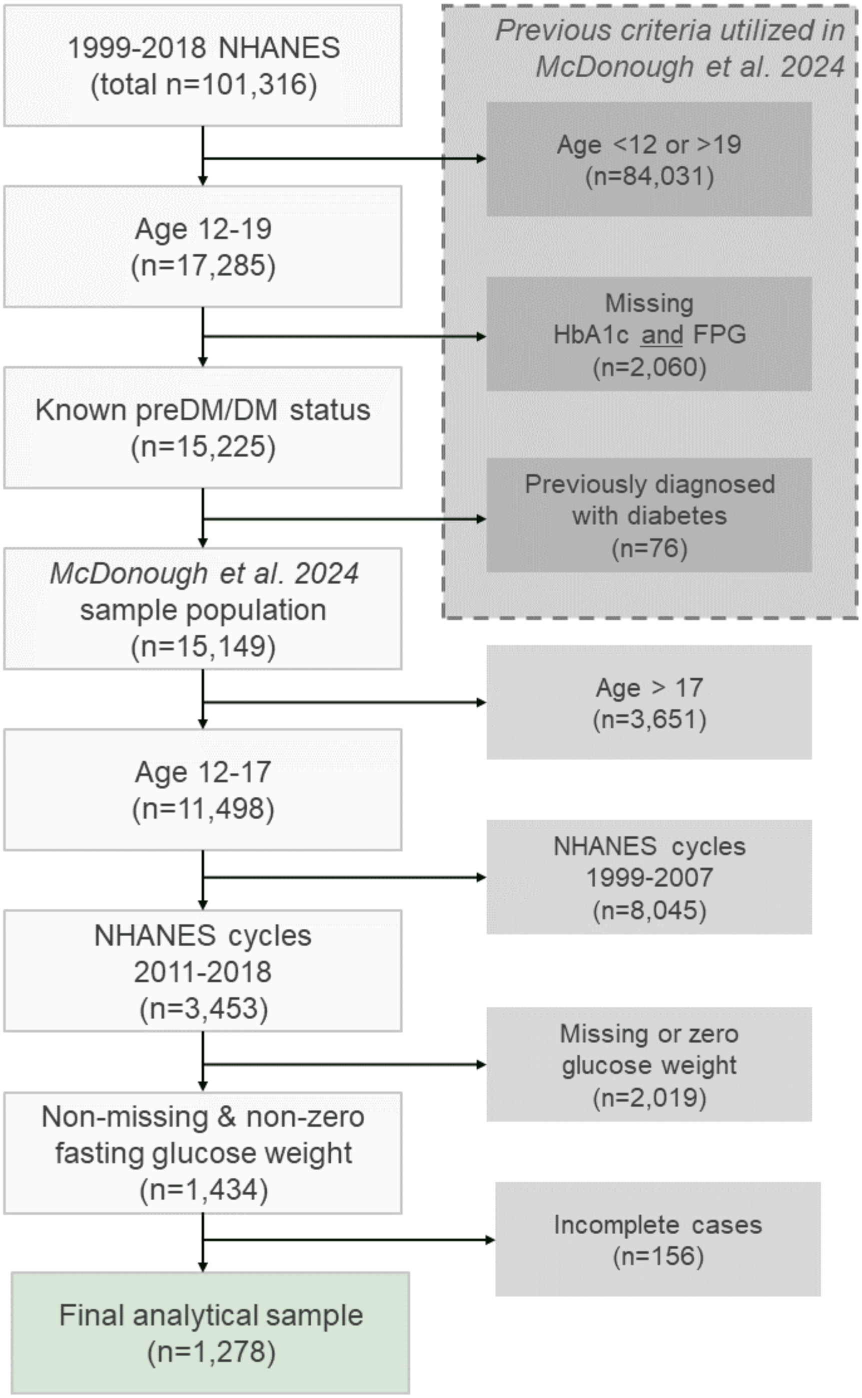

**Figure S2. Bootstrap indicating four classes as optimal solution for the latent class analysis**

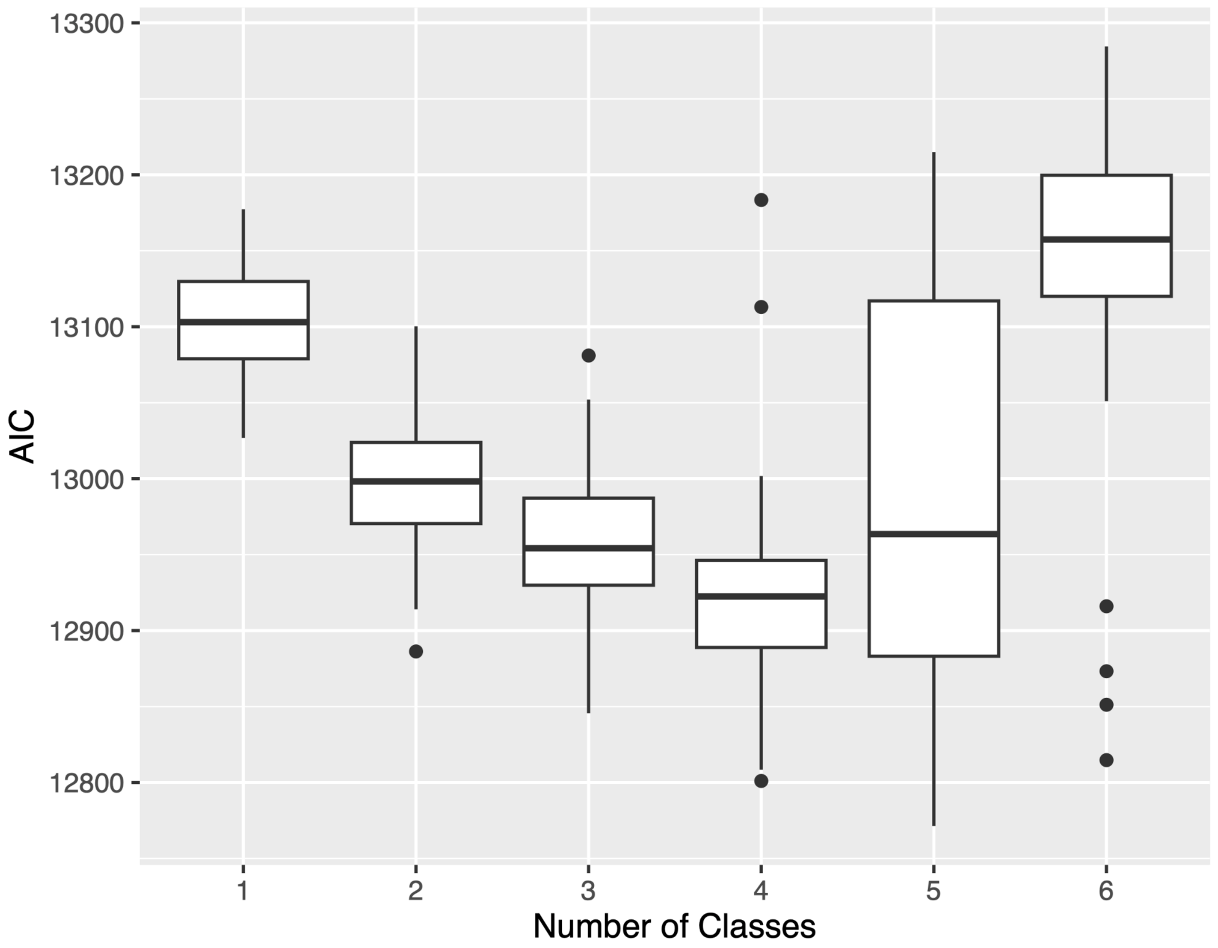

**Figure S3. Class-specific probabilities of BMI and lifestyle behavior categories across four latent classes identified based on a 3-step approach based on methods by the Bolck, Croon, and Hagenaars (BCH).**

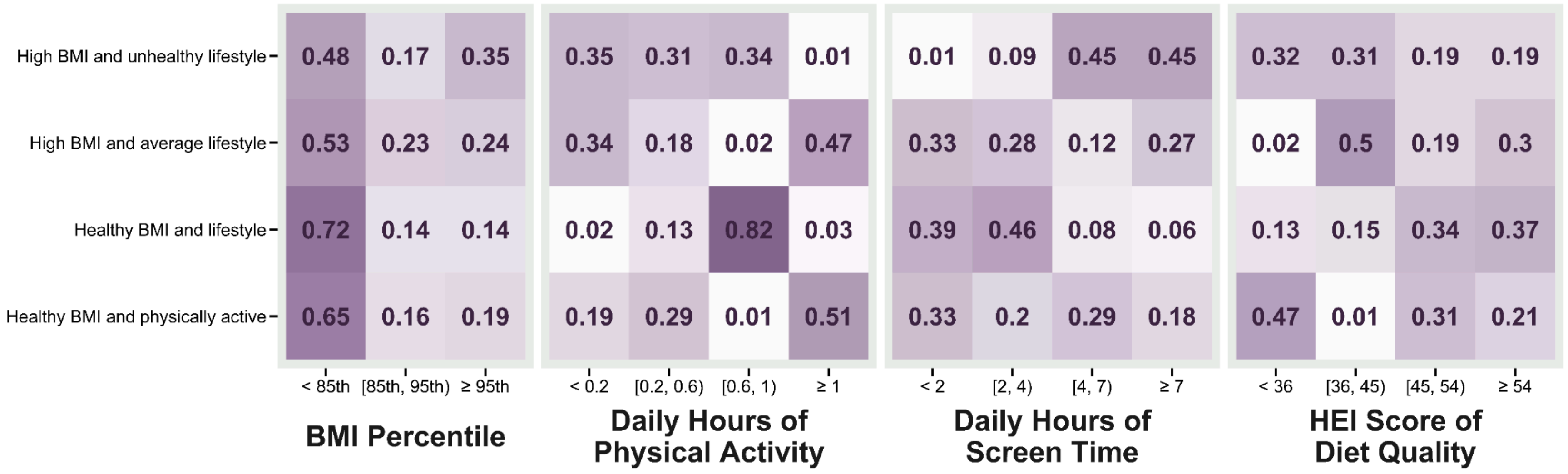


**Note**: We used an established SAS macro to implement the 3-step BCH-approach to identify latent classes and investigate the association between preDM/DM risk and the class membership. This 3-step approach differed from the main LCA analysis, which is a one-step approach where the coefficients on the covariates are estimated simultaneously *(48)*. as part of the latent class model. The BCH-approach included the following 3 steps: 1) identify the latent class without the covariates; 2) compute a special weighting variable using the posterior probabilities of class membership based on the LCA model; and 3) estimating multinomial logistic regression coefficients for predicting membership in each class with incorporation of the weighting variable from step-2. We tested the LCA model with 2 to 8 classes, and chose four classes as the optimal number of latent classes based on the minimum AIC associated with the 4-class solution. The four classes were similar to those identified in the main LCA model using the one-step approach, as shown in the class-specific probabilities (Figure S2). In the multinomial logistic regression, class membership was the outcome variable, where we chose the "High BMI and unhealthy lifestyle" as the reference class, and preDM/DM status was the exposure variable. The model additionally adjusted for age, sex, family PIR, insurance, as in the main model. NHANES survey weight was incorporated in the 3-step approach, but not clustering.

### Table S1. Raw NHANES variables and corresponding cycles used to build physical activity composite variable

| Cycle | Intensity | Frequency | Duration | Physical activity (PA) minutes per week | Physical activity (PA) hours per day |
|---|---|---|---|---|---|
| 2007-2016 | Moderate | **PAQ670**<br>Days moderate recreational activities *(per week)* | **PAD675**<br>*Minutes* moderate recreational activities | $Moderate\ PA\ min\ per\ week = PAQ670 \times PAD675$ | $PA\ hours\ per\ day = \frac{(Moderate\ PA\ min\ per\ wk + Vigorous\ PA\ min\ per\ wk)}{7\ days * 60\ minutes}$ |
| | Vigorous | **PAQ655**<br>Days vigorous recreational activities *(per week)* | **PAD660**<br>*Minutes* vigorous recreational activities | $Vigorous\ PA\ min\ per\ week = PAQ670 \times PAD675$ | |
| 2017-2018 | NA | **PAQ706**<br>Days physically active at least 60 min. | 60 minutes | | $PA\ hours\ per\ day\ * = \frac{PAQ706}{7\ days}$ |

* Based on the assumption that each day reported in PAQ706 is 60 minutes of physical activity.
